# Automating Individualized Patient Notification of Drug Recalls: Complex Challenges

**DOI:** 10.1101/2024.09.18.24312141

**Authors:** Meghana D. Gadgil, Rose Pavlakos, Simona Carini, Brian Turner, Ileana Elder, William Hess, Lisa Houle, Lavonia Huff, Elaine Johanson, Carole Jones, Daphne Liang, Pamela Ogonowski, Joshua Phipps, Tyler Peryea, Ida Sim

## Abstract

**Background:** Consumer-level drug recalls usually require action by individual patients. FDA has public-facing outlets to inform consumers and healthcare professionals about drug safety information including recalls, but consumers may not be aware of them. Prescribers are not notified about which of their patients are affected. The process may lead some patients to stop taking their medications. <u>Aim</u> To leverage the FDA’s Healthy Citizen prototype platform, which provides information about recalls, to automatically notify patients of relevant recalls.

**Method:** We developed and evaluated an electronic notification system in the Primary Care and Cardiology practices at a large urban, academic medical center. Using structured interviews, we assessed qualitative feedback on the system and portal messaging from a convenience sample of 9 patients.

**Results:** <u>Program Description</u> The healthcare portal scanned the FDA Healthy Citizen Application Programming Interface nightly to detect new recalls, identified patients who had those medications in their electronic health record (EHR) medication list, and sent them a message through the EHR patient portal with a link to a customized FDA information display. <u>Program Evaluation</u> The system was technically functional, but it was not possible to trace a medication prescription from the EHR to specific lot numbers dispensed to that patient by a community pharmacy.

**Discussion:** Lack of an accurate electronic audit trail from prescription to dispensed medication precludes clinical deployment of automated drug recall notification.

## INTRODUCTION

In the United States, the Food and Drug Administration (FDA) is responsible for assuring the safety and efficacy of marketed drugs. When a safety concern arises on a marketed drug, communicating this information to patients is essential and timely clinical action by prescribers is often required. Yet, patients and prescribers often lack relevant timely information, leaving patients and health systems unable to efficiently manage drug recalls and their impacts. Recognizing this problem, the FDA developed prototype technology for patients and health systems to automatically be notified of drug recalls through their healthcare portals, as part of the FDA’s Healthy Citizen prototype platform that seeks to allow “citizens and those who care for them, research organizations, and FDA to communicate and collaborate in a single, seamless environment connected through the healthcare portal and leveraging the trusted relationships between providers and patients to improve public health outcomes” (1). With FDA funding through the UCSF-Stanford Center of Excellence in Regulatory Science and Innovation (CERSI) (2), the University of California, San Francisco (UCSF), an academic medical center, partnered with the FDA to demonstrate use of Healthy Citizen tools to automate individualized drug recall notifications to outpatient Primary Care and Cardiology patients.

### Drug Recall Process

Firms, including manufacturers and own-label distributors, can initiate a recall, either on their own or in response to an FDA recommendation, request, or order. Common reasons for recalls include contamination, mislabeling, adverse reaction, defective product, and incorrect potency (3). The FDA works with firms as they develop their recall strategy, which is dependent on a variety of factors, including, but not limited to, the product’s degree of hazard, ease in identifying product, and the extent of distribution. Depth of recall is one component of this strategy: consumer-level recalls should be extended to consumers/patients; retail-level recalls affect community pharmacies and healthcare providers; and wholesale-level recalls affect manufacturers and distributors.

For consumer-level recalls, which were the focus of this project, consumers may learn of a recall through FDA.gov (4), news media, or notification from the recalling firm or pharmacy. Consumer notifications often recommend that patients consult their healthcare provider about the best course of action. However, recalls often affect only certain lots of pills and prescribers have no way of knowing the lot number of the medication dispensed to the patient and therefore whether the patient is affected. The patient often cannot identify the lot number either: dispensing pharmacies are not required to document the lot number on pill bottle labels. Thus, if patients contact their providers about a recall, the only action providers can take is to redirect patients to their pharmacy. The pharmacy then either replaces the pills with those from an unaffected lot or, if no substitution is available, notifies the prescribing clinician to issue a new prescription for a different medication, dosage, or formulation.

This partnership with the FDA aimed to address the inefficiencies in recall notification by demonstrating timely, fully automated, and individualized communication of drug recalls and recommended actions to patients.

## SETTING AND PARTICIPANTS

We developed an electronic notification system and conducted this study in the General Internal Medicine and Cardiology clinics at a large urban academic medical center in San Francisco, CA. The Primary Care clinic serves 25,000 patients with approximately 70,000 visits yearly. The Cardiology clinics serve over 12,000 patients with approximately 30,000 visits yearly. The clinics use the Epic® EHR with the MyChart patient portal. At the time of the study, approximately 45% of patients had actively used MyChart at least once.

We created and tested the notification system within Epic’s ACE6 development environment, with the intent to migrate it to production after successful testing. The project team was comprised of clinicians and programmers from the medical center, FDA leaders from Health Informatics and other sections, and developers of the Healthy Citizen platform. For testing purposes, fictitious patients were created within the Epic® ACE6 environment with medication lists that contained prescriptions matching fictitious medication recalls issued by the FDA.

The prototype system was shown to a convenience sample of 9 patients via remote videoconferencing to obtain initial formative feedback. This program evaluation was approved by the UCSF Institutional Review Board.

## PROGRAM DESCRIPTION

The notification system comprised two major technical parts. The first part, within the medical center’s firewalls, checked for new consumer-level drug recalls and notified affected patients via MyChart. The second part was the FDA’s Healthy Citizen prototype platform, which provided an Application Programming Interface (API) for external systems to request the latest drug recall information and mechanisms to launch a widget displaying details about a specific recall. The widget was a SMART-on-FHIR software module that could be embedded into and accessed within an EHR without the need for any additional sign-in.

### EHR build

The EHR build had three major parts: 1) Checking for new drug recalls; 2) Matching recalls the patient medication lists; and 3) Preparing and sending personalized MyChart notifications. Each part proved extremely challenging to build for technical and data availability reasons.

First, the system issued a nightly call to the Healthy Citizen API to retrieve the National Drug Codes (NDC) of newly recalled drugs. The next step, matching recalls to a patient’s EHR medication list, can result in false negatives and false positives. False negatives can occur if a patient’s prescription is missing from the medication list (5), or if the algorithm fails detects a true match. False positives can arise from two inaccuracies. Crucially, EHR medication lists contain the *prescribed* drug, not the *dispensed* drug. To identify a prescribed drug, Epic uses RxNORM codes that do not include the manufacturer name. To identify recalled drugs and their manufacturers, the FDA uses NDC codes, which are unique, three-segment numbers that identify a drug’s labeler (i.e., manufacturer or distributor), product, and trade package size (6). Thus, for example, the NDC from a lisinopril recall from a specific manufacturer will match the RxNORM code for all lisinopril prescriptions of the same strength, regardless of manufacturer.

This will erroneously identify patients who were prescribed lisinopril but were not dispensed pills from the affected manufacturer. Secondly, recalls often involve only specific lots, information unavailable in the EHR thus contributing to false positives as discussed above.

The third part of the EHR build was to send a MyChart notification to patients once a match was made, alerting them they may be taking a recalled medication (Figure 1).

**Figure 1.**
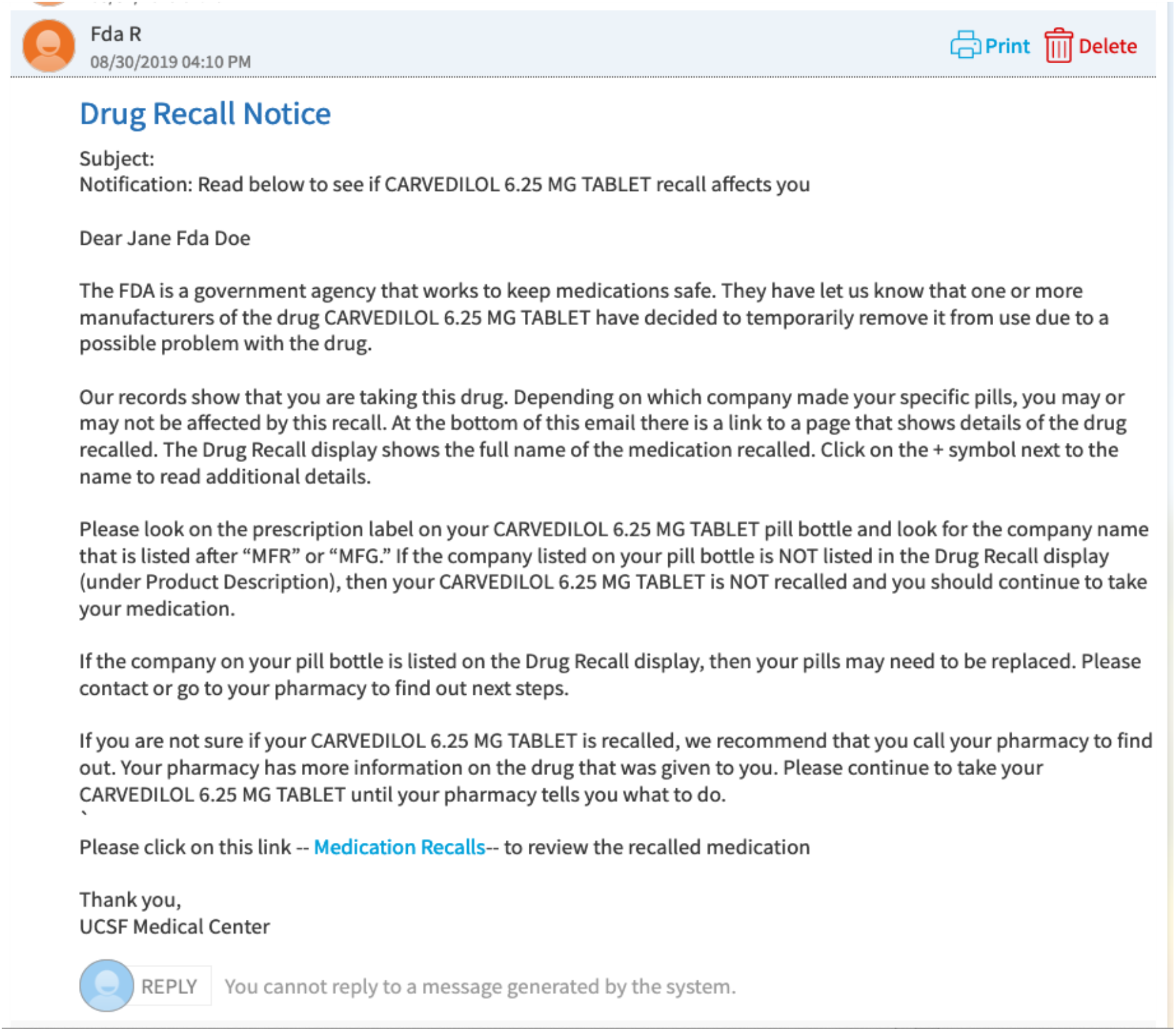
MyChart notification of potentially relevant recall.

The link to the FDA display launched a new window within MyChart showing details of the matched recall, including affected manufacturers (Figure 2). Because the matching algorithm could not restrict matches to affected manufacturers, the MyChart message asked patients to compare the manufacturer name on their pill bottle’s label to the manufacturer(s) listed in the FDA informational display and to call their pharmacy if it matched. The Patient Advisory Council of the Primary Care clinic provided input on the wording and endorsed the importance of the project aims.

**Figure 2.**
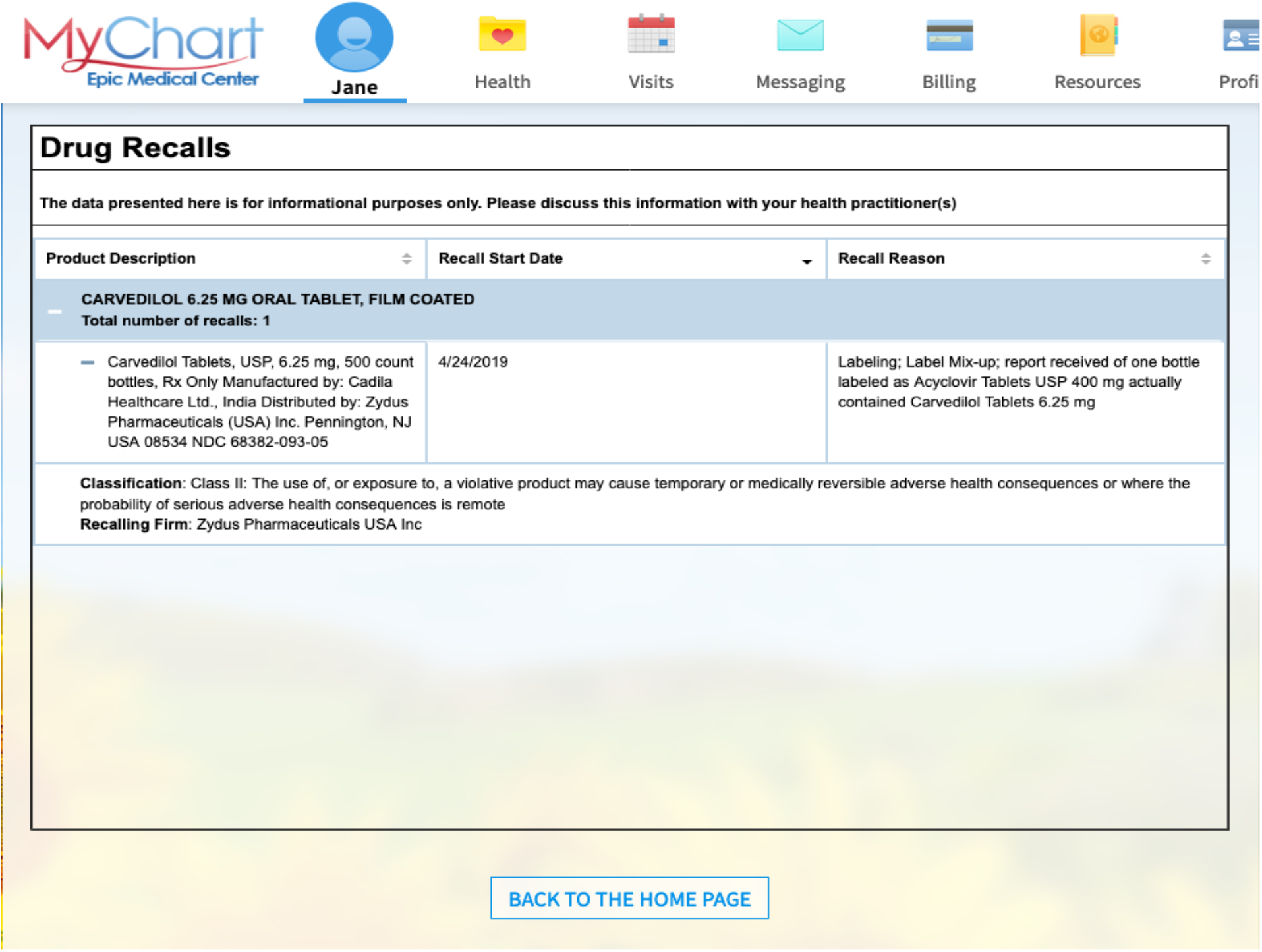
FDA Health Citizen information display widget showing official information about a drug recall.

### Healthy Citizen build

Substantial technical work was performed on the Healthy Citizen platform to satisfy the use case needs. For example, technical and internal FDA administrative changes were required to make the depth of recall (i.e., retail- or consumer-level) searchable, and to distinguish between new and ongoing recalls. The SMART-on-FHIR widget needed to be available on Epic’s App Orchard, and modifications were required for the widget to be called by and launched within Epic. The contents of the widget display were modified to exclude information not relevant to patients such as status of recall (e.g., ongoing or completed) or move it from the main display to the additional details section.

## PROGRAM EVALUATION

The system was able to automatically detect a new fictitious medication recall using the Healthy Citizen API, compare and detect matches to each (fictitious) patient’s list of prescribed medications, send a MyChart message to affected (fictitious) patients, and launch a display for the correct recall(s). The system responded correctly to test patients with none to multiple affected medications.

We obtained qualitative feedback by interviewing a convenience sample of 9 patients (5 female, 4 male), all MyChart users, recruited via email from the Primary Care and Cardiology Clinics. Two of the 9 participants had personal experience with recalls. Participants were interviewed individually using Zoom. During the session, they were presented with a scenario for fictitious patient Jane Doe, who was prescribed carvedilol 6.25 mg. Using structured interviewing techniques, we evaluated participants’ understanding of the MyChart message, widget, and example pill bottle label. Throughout the process we asked for descriptive feedback. Recordings of the interviews were transcribed and separately analyzed by two investigators (MG, RP) for common themes, with additional verification by SC and IS.

All 9 participants understood the purpose of the MyChart notification message but thought it was too wordy. All 9 were able to identify the medication manufacturer on the example pill bottle label. Only 2 would have clicked on their own on the link at the bottom of the MyChart message to launch the widget; the other 7 needed the interviewer’s prompting and guidance to do so. As advised by the MyChart message, all 9 users would have contacted their pharmacist, but 5 of the 9 would also have contacted their doctor’s office as advised by the widget. Given the choice, all 9 would have liked to receive MyChart notification of potential drug recalls.

Major thematic findings included the following. 1) Patients appreciated being notified of recalls by their clinic even though their actual medication may not have been affected by the recall, because they trusted the clinic and the notification showed that the clinic was aware of patient medication issues. 2) Communicating through the MyChart patient portal was seen as a trusted, efficient, and reliable notification method. Mailed letters can be ignored, and several users said they did not answer phone calls from unknown phone numbers (e.g., their pharmacy). 3) The widget content should be displayed directly in the MyChart message rather than in a new window. 4) The widget itself should be re-designed to more directly meet patient information needs. Much of the widget content was either confusing or irrelevant to patients (e.g., Recall

Start Date, manufacturer address). The Recall Reason was appreciated but was felt to be unnecessary. The widget should not ask patients to discuss the information with their healthcare provider. 5)Patients wanted to discuss the recall with their clinicians to “close the loop.”

The project team concluded that operational deployment of this system may lead to unnecessary and unacceptable patient anxiety generated by false positive notifications. In addition, because patient feedback suggested that patients would contact their clinicians regardless of the advice to contact their pharmacy, the system was likely to increase staff burden for responding to patient inquiries. We therefore decided not to proceed with implementation of the FDA drug recall notification system into clinical care.

## DISCUSSION

Patients and clinicians need an accurate system for identifying which patients are affected by which drug recalls and acting on them in a timely and appropriate manner to prevent patient harm and erosion of trust in prescribers and the healthcare system.

This project demonstrated the technical and clinical feasibility of using the FDA’s Healthy Citizen drug recall tools to automatically alert patients, via Epic’s patient portal MyChart, to relevant drug recalls. While our project was technically successful, it revealed substantial challenges to responding to drug recalls. Chiefly, while patients want and expect their prescriber to be aware of, and involved in, responding to a drug recall, prescribers have no easy access to the manufacturer and lot number of the actual medication dispensed to their patients. Without these details, health systems cannot accurately target patients and false positive notifications are inevitable. A partial technical solution could be to access Surescripts records that include the NDC for dispensed drugs as reported to Surescripts via claims data. However, only 70% of patients at USCF medical center use a Surescripts-participating pharmacy and Surescripts records do not include dispensed lot numbers, such that false positive recall notifications would still be an issue.

Our project showed that a strong case can be made for requiring each pill bottle to include on its label the lot number and NDC code of the pills (which links to the manufacturer, labeler, or distributor), so that patients could definitively determine if a recall affected them. Current federal regulation allows such information to appear on an internal leaflet or a label on the outer carton or wrapper of manufactured medications (7), which many patients discard even if the pharmacist includes them with the dispensed medication. Only three State Boards of Pharmacy require the NDC to appear on the dispensed medication label, and only five State Boards of Pharmacy require the lot number to appear on the dispensed medication label. The manufacturer and lot number of dispensed medications should routinely be available electronically to prescribing clinicians via standard APIs so that health systems can meet patient expectations of being a trusted guide in properly responding to drug recalls. Policy and data infrastructure changes are required at the regulatory, health IT, and consumer pharmacy levels before automated recall notification can be widely deployed.

## Data Availability

Data produced in the present work are summarized in the manuscript

## Acknowledgements

This publication was made possible by Grant Number U01FD005978 from the FDA, which supports the UCSF-Stanford Center of Excellence in Regulatory Sciences and Innovation. Its contents are solely the responsibility of the authors and do not necessarily represent the official views of the HHS or FDA. This project was funded with $235,000 (representing 100%) by the FDA/HHS.

This work was presented at the Innovations in Regulatory Science Summit, January 2020 and at APhA2020, March 2020.

